# Predicting Wilson’s Disease Progression using Machine Learning with Real-World Electronic Health Records

**DOI:** 10.1101/2023.07.28.23293309

**Authors:** Caihua Liang, Scott P. Kelly, Rongjun Shen, Ling Li, Kasia Lobello, Steven Arkin, Kui Huang, Xiaofeng Zhou

**Author notes:** **Corresponding author:** Caihua Liang, MD, PhD, Global Medical Epidemiology, Worldwide Medical & Safety, Pfizer, Inc., 235 East 42nd Street, New York, NY 10017. **Author contributions** Concept and design: Liang, Kelly, Shen, Li, Lobello, Arkin, Huang, Zhou. Acquisition, analysis, or interpretation of data: Liang, Shen, Li, Zhou. Drafting of the manuscript: Liang. Revision of the manuscript: Liang, Li, Kui, Zhou. Statistical analysis: Liang, Shen, Ling. **Conflict of Interest Disclosures:** Liang, Kelly, Shen, Li, Lobello, Arkin, Huang, Zhou were Pfizer employees at the time of this manuscript preparation and are shareholders in Pfizer. **Funding/Support:** This study was sponsored by Pfizer.

## Abstract

**BACKGROUND & AIMS:** Wilson’s disease (WD) is a rare genetic disorder causing excessive copper accumulation. Research on the natural history of WD is limited. Our objective was to identify predictors for WD progression to cirrhosis, liver failure, and death and to predict individual risk of progression to these endpoints at 1, 2, 3, and 5 years after WD diagnosis.

**METHODS:** A retrospective natural history cohort study of adult patients with first-recorded WD diagnosis was conducted using the US Optum EHR data between 1/1/2007 and 6/30/2020. LASSO Cox regression, Random Survival Forest (RSF), and XGBoost (XGB) models were used to identify important predictors for progression to cirrhosis, liver failure, and death. The strong predictors for each outcome identified through weighted average rankings across models and reviewed by clinical experts were used for patient-level prediction using RSF and XGB models. The resulting models were validated with an independent sample cohort. C-index and dynamic AUCs were used to evaluate model performance.

**RESULTS:** Over the study period, 310 out of 2,901 WD patients developed cirrhosis, 255 out of 3,251 developed liver failure, and 604 out of 3,559 died. Age at WD diagnosis, alcoholism, AST and bilirubin levels within 3 months of WD diagnosis, and neurologic and hepatic conditions were the most common predictors for progression to the study endpoints. XGB had a slight superior predictive performance compared with RSF and was then used to predict individual risks for progression to the study endpoints with the top ensemble predictors. The dynamic AUC was 0.78 at Year 1, 0.74 at Year 2, 0.72 at Year 3 and 0.72 at Year 5 for cirrhosis; 0.82 at Year 1, 0.78 at Year 2, and 0.77 at both Year 3 and Year 5 for liver failure; 0.81 at Year 1, 0.83 at Year 2, and 0.82 at both Year 3 and Year 5 for death.

**CONCLUSIONS:** This study identified the most influential clinical predictors and assessed patient-level risk of WD progression using machine learning. Results from machine learning prognostic models will increase understanding of disease natural history and may help improve clinical trial design and guide individualized clinical care.

---

Wilson’s disease (WD) is a rare genetic disorder caused by the mutations in the copper-transporting gene, ATP7B.^1^ Variations of ATP7B causes excessive copper accumulation, particularly in liver, brain, and eyes. Accumulation of free copper in the liver induces hepatocyte dysfunction, which may initially manifest as steatosis and later progress to hepatitis, cirrhosis, fibrosis, liver failure, or death.^2^

It was estimated that the prevalence of WD was 1 case per 30,000 live births in most populations.^2^An estimate of 40-50% of WD will present with symptomatic hepatic diseases, but manifestations range from mild hepatic dysfunction to liver failure in a broad spectrum.^3^ In the absence of straightforward diagnosis or curative treatment for WD, it is essential to understand its natural history and progression to advanced stages. Identification of risk factors, prediction of WD progression, and understanding which patient subgroup(s) may benefit from a particular treatment are useful for clinical trials design and for patients to make optimal clinical care decisions.

A machine learning technique enables identifying important features and modeling disease progression using real-world data. Machine learning techniques with survival analysis automatically incorporate a large array of features in a nonlinear pattern and use multiple interactions to effectively improve the performance of traditional proportional hazard models in identifying critical features.^4–6^ RSF, an ensemble tree method developed by Ishwaran H et al in 2008,^7^ is one of the most efficient models in survival analysis because of easy parameter turning, high data adaptability and absence of model assumptions.^8^ XGB, a novel and complex ensemble algorithm published in 2016^9^, is widely recognized because of its scalability and fast learning. XGB also combines the advantages of bagging and boosting methods and effectively improve the accuracy of prediction.^10^

In this study, we developed an exploratory framework using machine learning that aims at predicting WD progression. Two steps are involved with first to identify important predictors for WD progression to cirrhosis, liver failure, and death and second to use these important predictors to predict individual risk of progression to these outcomes at 1, 2, 3, and 5 years after WD diagnosis.

## Materials and Methods

### Data Source

The patients in this study were identified from Optum’s EHR Database derived from the electronic health records of a network of healthcare provider organizations across the United States that include more than 700 hospitals and 7000 clinics. This database incorporates clinical and medical administrative data from both inpatient and ambulatory EMRs, practice management systems, and numerous other internal systems. The data are processed from across the continuum of care, including acute inpatient stays and outpatient visits. Only the medical records from the Integrated Delivery Networks (IDNs) were used in this study to better capture patients’ continuum of care by linking inpatient and outpatient data.

### Study Design and Study Population

This was a retrospective cohort study of WD natural history between January 1, 2007 and September 30, 2020. Study population consisted of individuals aged 18 years over with a WD diagnosis code (ICD 10 code: E83.01 or ICD 9 code: 275.1) associated with at least one inpatient visit or at least two outpatient visits (at least 30 days apart). Eligible patients were required to have at least 12 months baseline period prior to the first-recorded WD diagnosis and were excluded for any prior WD diagnosis or any prior events of interest during the baseline period. Patients were followed from the first-recorded WD diagnosis (index date) to the earliest date of first occurrence of each study event, end of database observation, and end of study period (September 30, 2020). Potential predictors and WD features were collected in terms of demographics, comorbidities, treatments, lab results and health care utilization.

This study using deidentified structured data from a secondary database and, hence, independent ethics committee and/or institutional review board review was not required.

### Outcome Measures

We assessed patient’s progression from WD diagnosis to cirrhosis, liver failure and all-cause of death. Cirrhosis and liver failure were identified using diagnosis codes; all-cause of death was determined via linkage to the Social Security Administration’s Death Master File or as indicated within the medical record.

## Statistical Analysis

### Data pre-processing

There were a wide range of potential predictors (∼180) for selection on a priori hypothesis based on the literature and clinical knowledge, including demographics, comorbidities, treatments, lab results and health care utilization as well as the 30 most frequent diagnoses, procedures, and medications. Detailed data pre-processing is provided in the supplemental materials.

### Ensemble Variable Selection

Feature selection for each outcome was performed using least absolute shrinkage and selection operator (LASSO) regression, RSF, and XGBoost (XGB) models (Detailed model parameters are provided in the supplemental materials). The importance of the candidate predictors associated with cirrhosis, liver failure, and death respectively was ranked based on the values of absolute coefficients from LASSO, variable importance from RSF, and gain information from XGB. Given the potential bias resulting from a single feature section method, an ensemble method using weighted average ranking was applied to incorporate the results across the three models and to quantify the ensemble importance in an overall ranking of the important predictors for each outcome.^14^ The top 50 resulting predictors from the ensemble selection for each outcome were reviewed by clinical experts for biological plausibility. The final predictors (17 features for cirrhosis, 16 for liver failure, and 26 for death) considered to be statistically and clinically meaningful were used for patient-level progression prediction.

### Predicting Models

The time-to-event analysis of RSF and XGB was leveraged to predict the individual risk of developing cirrhosis, liver failure, and death within time windows of 1 year, 2 years, 3 years, and 5 years from diagnosis of WD (detailed modeling process is provided in the supplemental material). A random sample of 70% of the study cohort were selected for training the model and the 30% holdout set were used for validation. Harrell’s concordance index (C-index) was calculated to evaluate the model performance in the validation set.^7^ C-index, a global measure of discriminative power of a survival model, is defined as the fraction of pairs of patients who have a longer survival time are also predicted with lower risk score by the model. The model with a better performance will be used to predict individual risks of outcomes. Area under the receiver operating characteristics curve (AUC) was also calculated, which measures the probability that a higher score was assigned to a random positive outcome than a random negative one. Dynamic AUCs at Year 1, Year 2, Year 3, and Year 5 were generated and illustrated. All analyses were performed using R Studio with packages specified in the supplemental materials.

### Data Availability

The data that support the findings of this study are available from Optum but restrictions apply to the availability of these data, which were used under license for the current study, and so are not publicly available. Data are however available from the authors upon reasonable request and with permission of Optum.

## Results

### Baseline Characteristics

Table 1 shows the descriptive distribution of select baseline covariates by study cohorts and outcomes of interest. In the cirrhosis cohort, 2,901 patients were diagnosed with WD, of whom 310 developed cirrhosis. Compared with patients without cirrhosis, cirrhosis patients had more males (54.8% vs 41.1%); had a higher proportion of having chronic hepatitis (12.6% vs 1.9%), alcoholism (15.2% vs 4.4%), acute hepatitis (9.4% vs 1.7%), presence of both neurologic and hepatic conditions (14.2% vs 8.8%), hepatic steatosis (19.7% vs 7.7%), diabetes (28.1% vs 17.9%), obesity (21.0% vs 16.0%), and coronary artery disease (19.0% vs 11.2%); and were more likely to have elevated bilirubin levels (12.6% vs 4.6%) and AST>=70 U/L (15.5% vs 5.2%).

**Table 1.** Descriptive Distribution of Select Baseline Covariates by Study Cohorts and Outcomes of Interest.

| Characteristics | Cirrhosis |  |  | Liver Failure |  |  | Death |  |  |
| --- | --- | --- | --- | --- | --- | --- | --- | --- | --- |
|  | Yes | No | HR (95 %CI) | Yes | No | HR (95 %CI) | Yes | No | HR (95 %CI) |
| Total N | 310 | 2,591 |  | 255 | 2,996 |  | 604 | 2,955 |  |
| **Demographics** |  |  |  |  |  |  |  |  |  |
| Age | 49.6 ± 17.3 | 48.7 ± 19.9 | 1.0(1.0-1.0) | 50.4 ± 16.0 | 49.4 ± 19.5 | 1.0(1.0-1.0) | 62.1 ± 15.4 | 46.7 ± 18.7 | 1.0(1.0-1.1) |
| Gender (Reference level: Female) |  |  |  |  |  |  |  |  |  |
| Male | 170(54.8%) | 1,066(41.1%) | 1.7(1.4-2.1) | 119(46.7%) | 1,308(43.7%) | 1.1(0.9-1.5) | 304(50.3%) | 1,265(42.8%) | 1.3(1.1-1.5) |
| **Medical History/Comorbidities** |  |  |  |  |  |  |  |  |  |
| Chronic hepatitis | 39(12.6%) | 49(1.9%) | 6.3(4.5-8.8) | 32(12.5%) | 132(4.4%) | 3.1(2.1-4.5) | 43(7.1%) | 164(5.5%) | 1.4(1.1-2.0) |
| Alcoholism | 47(15.2%) | 114(4.4%) | 3.6(2.7-5.0) | 71(27.8%) | 182(6.1%) | 5.7(4.4-7.6) | 102(16.9%) | 285(9.6%) | 2.2(1.8-2.7) |
| Acute hepatitis | 29(9.4%) | 45(1.7%) | 5.3(3.6-7.7) | 18(7.1%) | 105(3.5%) | 2.1(1.3-3.5) | 34(5.6%) | 135(4.6%) | 1.4(1.0-2.0) |
| Neurologic or hepatic conditions (Reference level: No neurologic or hepatic conditions) |  |  |  |  |  |  |  |  |  |
| ≥ 1 hepatic condition but no neurologic conditions | 100(32.3%) | 397(15.3%) | 2.9(2.2-3.7) | 124(48.6%) | 605(20.2%) | 5.7(4.1-7.8) | 157(26.0%) | 766(25.9%) | 1.6(1.3-2.0) |
| ≥ 1 neurologic condition but no hepatic conditions | 36(11.6%) | 575(22.2%) | 0.8(0.5-1.1) | 20(7.8%) | 591(19.7%) | 1.0(0.6-1.7) | 110(18.2%) | 501(17.0%) | 1.6(1.3-2.1) |
| ≥ 1 neurologic condition and ≥ 1 hepatic conditions | 44(14.2%) | 227(8.8%) | 2.6(1.8-3.6) | 56(22.0%) | 333(11.1%) | 5.3(3.7-7.7) | 129(21.4%) | 374(12.7%) | 3.1(2.5-3.8) |
| Elevation of lactic acid dehydrogenase level | 76(24.5%) | 300(11.6%) | 2.7(2.1-3.5) | 53(20.8%) | 406(13.6%) | 1.8(1.3-2.4) | 89(14.7%) | 459(15.5%) | 1.2(0.9-1.5) |
| Hepatic steatosis | 61(19.7%) | 200(7.7%) | 3.0(2.3-4.0) | 49(19.2%) | 314(10.5%) | 2.2(1.6-3.0) | 69(11.4%) | 393(13.3%) | 1.1(0.9-1.4) |
| Diabetes | 87(28.1%) | 464(17.9%) | 1.8(1.4-2.3) | 79(31.0%) | 603(20.1%) | 1.8(1.4-2.4) | 216(35.8%) | 554(18.7%) | 2.2(1.9-2.6) |
| Obesity | 65(21.0%) | 414(16.0%) | 1.5(1.2-2.0) | 63(24.7%) | 524(17.5%) | 1.7(1.3-2.2) | 113(18.7%) | 570(19.3%) | 1.2(1.0-1.4) |
| Kayser-Fleischer rings | 7(2.3%) | 11(0.4%) | 4.0(1.9-8.5) | 6(2.4%) | 18(0.6%) | 3.7(1.7-8.4) | 2(0.3%) | 31(1.0%) | 0.3(0.1-1.4) |
| Choreiform movements | 0(0.0%) | 14(0.5%) | 0.0(0.0-0.0) | 1(0.4%) | 14(0.5%) | 1.1(0.2-7.8) | 5(0.8%) | 14(0.5%) | 2.3(1.0-5.6) |
| Coronary artery disease | 59(19.0%) | 289(11.2%) | 1.9(1.5-2.6) | 29(11.4%) | 379(12.7%) | 1.0(0.7-1.4) | 167(27.6%) | 300(10.2%) | 2.9(2.4-3.5) |
| Cirrhosis | 0(0.0%) | 0(0.0%) |  | 113(44.3%) | 215(7.2%) | 9.6(7.5-12.3) | 130(21.5%) | 380(12.9%) | 2.2(1.8-2.7) |
| Ascites | 32(10.3%) | 87(3.4%) | 3.4(2.4-4.9) | 81(31.8%) | 164(5.5%) | 7.8(6.0-10.2) | 130(21.5%) | 306(10.4%) | 2.8(2.3-3.4) |
| Anemia | 118(38.1%) | 908(35.0%) | 1.2(1.0-1.5) | 140(54.9%) | 1,048(35.0%) | 2.3(1.8-3.0) | 346(57.3%) | 1,034(35.0%) | 2.5(2.1-2.9) |
| Parkinson's disease | 8(2.6%) | 45(1.7%) | 1.7(0.8-3.4) | 5(2.0%) | 51(1.7%) | 1.3(0.5-3.1) | 14(2.3%) | 53(1.8%) | 1.6(0.9-2.7) |
| Liver fibrosis | 4(1.3%) | 14(0.5%) | 3.8(1.4-10.1) | 82(32.2%) | 175(5.8%) | 6.7(5.2-8.8) | 100(16.6%) | 279(9.4%) | 1.9(1.6-2.4) |
| Psychosis | 11(3.5%) | 104(4.0%) | 0.9(0.5-1.7) | 21(8.2%) | 119(4.0%) | 2.2(1.4-3.5) | 44(7.3%) | 125(4.2%) | 1.9(1.4-2.6) |
| Personality disorders | 4(1.3%) | 47(1.8%) | 0.9(0.3-2.4) | 9(3.5%) | 48(1.6%) | 2.6(1.4-5.2) | 10(1.7%) | 56(1.9%) | 1.4(0.7-2.6) |
| Renal dysfunction | 59(19.0%) | 442(17.1%) | 1.3(1.0-1.7) | 65(25.5%) | 527(17.6%) | 1.8(1.4-2.4) | 225(37.3%) | 515(17.4%) | 3.1(2.6-3.6) |
| Disorders of fluid, electrolyte, and acid-base balance | 63(20.3%) | 464(17.9%) | 1.3(1.0-1.7) | 87(34.1%) | 528(17.6%) | 2.5(1.9-3.3) | 239(39.6%) | 526(17.8%) | 3.0(2.6-3.5) |
| Cancer (except non melanoma skin cancer) | 28(9.0%) | 299(11.5%) | 0.8(0.6-1.2) | 26(10.2%) | 345(11.5%) | 1.0(0.6-1.4) | 139(23.0%) | 263(8.9%) | 2.7(2.2-3.3) |
| Renal/kidney failure | 47(15.2%) | 343(13.2%) | 1.3(1.0-1.8) | 52(20.4%) | 406(13.6%) | 1.8(1.3-2.4) | 200(33.1%) | 380(12.9%) | 3.3(2.8-3.9) |
| Hepatic encephalopathy | 0(0.0%) | 0(0.0%) |  | 0(0.0%) | 0(0.0%) |  | 58(9.6%) | 150(5.1%) | 2.6(2.0-3.5) |
| Hypertension | 143(46.1%) | 998(38.5%) | 1.4(1.1-1.7) | 124(48.6%) | 1,213(40.5%) | 1.4(1.1-1.8) | 385(63.7%) | 1,103(37.3%) | 2.7(2.3-3.1) |
| Gait disturbance | 34(11.0%) | 281(10.8%) | 1.2(0.8-1.7) | 37(14.5%) | 323(10.8%) | 1.6(1.1-2.3) | 105(17.4%) | 301(10.2%) | 2.2(1.7-2.7) |
| Dysphagia | 14(4.5%) | 162(6.3%) | 0.8(0.5-1.4) | 17(6.7%) | 191(6.4%) | 1.2(0.7-1.9) | 61(10.1%) | 179(6.1%) | 2.0(1.5-2.6) |
| Cardiovascular disease | 74(23.9%) | 498(19.2%) | 1.4(1.1-1.8) | 55(21.6%) | 592(19.8%) | 1.2(0.9-1.6) | 230(38.1%) | 511(17.3%) | 2.6(2.2-3.1) |
| Chronic kidney disease | 70(22.6%) | 473(18.3%) | 1.4(1.1-1.8) | 70(27.5%) | 563(18.8%) | 1.7(1.3-2.3) | 230(38.1%) | 539(18.2%) | 2.7(2.3-3.2) |
| **Lab results** |  |  |  |  |  |  |  |  |  |
| Bilirubin level (Reference level: 0.3-1.2 MG/DL) |  |  |  |  |  |  |  |  |  |
| <0.3 | 15(4.8%) | 247(9.5%) | 0.6(0.3-1.0) | 18(7.1%) | 261(8.7%) | 1.1(0.6-1.8) | 47(7.8%) | 240(8.1%) | 0.8(0.6-1.1) |
| >1.2 | 39(12.6%) | 119(4.6%) | 2.9(2.0-4.2) | 69(27.1%) | 163(5.4%) | 6.2(4.4-8.6) | 99(16.4%) | 259(8.8%) | 1.7(1.4-2.2) |
| Missing | 143(46.1%) | 1,224(47.2%) | 1.0(0.7-1.2) | 95(37.3%) | 1,408(47.0%) | 1.0(0.7-1.3) | 201(33.3%) | 1,406(47.6%) | 0.6(0.5-0.7) |
| AST level (Reference level: normal [$<35$ U/L]) | | | | | | | | | |
| Greater than normal to 2x upper limit [$35-<70$ U/L] | 57(18.4%) | 250(9.6%) | 3.4(2.4-4.8) | 59(23.1%) | 331(11.0%) | 3.9(2.7-5.8) | 99(16.4%) | 353(11.9%) | 1.2(0.9-1.5) |
| $\geq 2$x upper limit $\geq 70$ U/L] | 48(15.5%) | 135(5.2%) | 5.8(4.0-8.5) | 56(22.0%) | 185(6.2%) | 7.3(4.9-10.8) | 73(12.1%) | 254(8.6%) | 1.4(1.1-1.8) |
| Missing | 143(46.1%) | 1,198(46.2%) | 1.8(1.3-2.4) | 94(36.9%) | 1,380(46.1%) | 1.5(1.1-2.1) | 193(32.0%) | 1,385(46.9%) | 0.6(0.5-0.7) |
| ALT level (Reference level: normal [$<35$ U/L]) | | | | | | | | | |
| Greater than normal to 2x upper limit [$35-<70$ U/L] | 48(15.5%) | 285(11.0%) | 2.1(1.4-3.0) | 56(22.0%) | 350(11.7%) | 2.1(1.5-3.0) | 89(14.7%) | 364(12.3%) | 0.9(0.7-1.1) |
| $\geq 2$x upper limit $\geq 70$ U/L] | 47(15.2%) | 184(7.1%) | 3.2(2.2-4.6) | 31(12.2%) | 237(7.9%) | 1.8(1.2-2.7) | 49(8.1%) | 278(9.4%) | 0.7(0.5-0.9) |
| Missing | 144(46.5%) | 1,200(46.3%) | 1.4(1.1-1.9) | 94(36.9%) | 1,383(46.2%) | 0.9(0.6-1.2) | 196(32.5%) | 1,384(46.8%) | 0.5(0.4-0.6) |
| WBC level (Reference level: 4.5-11.0 *10<sup>9</sup> /L) |  |  |  |  |  |  |  |  |  |
| <4.5 | 34(11.0%) | 226(8.7%) | 1.6(1.1-2.3) | 51(20.0%) | 273(9.1%) | 2.5(1.8-3.5) | 78(12.9%) | 301(10.2%) | 1.3(1.0-1.6) |
| >11.0 | 13(4.2%) | 203(7.8%) | 0.5(0.3-1.0) | 17(6.7%) | 212(7.1%) | 1.0(0.6-1.6) | 100(16.6%) | 162(5.5%) | 1.8(1.4-2.2) |
| Missing | 155(50.0%) | 1,115(43.0%) | 1.2(1.0-1.6) | 96(37.6%) | 1,312(43.8%) | 0.9(0.7-1.2) | 166(27.5%) | 1,341(45.4%) | 0.5(0.4-0.6) |
| Creatinine level (Reference level: 0.7-1.3 MG/DL) |  |  |  |  |  |  |  |  |  |
| <0.7 | 52(16.8%) | 438(16.9%) | 1.1(0.8-1.5) | 53(20.8%) | 491(16.4%) | 1.3(0.9-1.8) | 100(16.6%) | 502(17.0%) | 0.8(0.6-1.0) |
| >1.3 | 20(6.5%) | 157(6.1%) | 1.3(0.8-2.2) | 23(9.0%) | 183(6.1%) | 1.7(1.1-2.6) | 102(16.9%) | 139(4.7%) | 2.6(2.0-3.2) |
| missing | 143(46.1%) | 1,076(41.5%) | 1.2(0.9-1.5) | 90(35.3%) | 1,264(42.2%) | 0.8(0.6-1.1) | 153(25.3%) | 1,296(43.9%) | 0.5(0.4-0.6) |
| Albumin level (Reference level: 35-55 G/L) |  |  |  |  |  |  |  |  |  |
| <35 | 53(17.1%) | 393(15.2%) | 1.2(0.9-1.7) | 98(38.4%) | 460(15.4%) | 3.9(2.8-5.3) | 229(37.9%) | 483(16.3%) | 2.8(2.3-3.4) |
| >55 | 0(0.0%) | 3(0.1%) | 0.0(0.0-0.0) | 0(0.0%) | 3(0.1%) | 0.0(0.0-0.0) | 0(0.0%) | 3(0.1%) | 0.0(0.0-0.0) |
| Missing | 144(46.5%) | 1,195(46.1%) | 1.0(0.8-1.3) | 94(36.9%) | 1,384(46.2%) | 1.2(0.8-1.6) | 202(33.4%) | 1,381(46.7%) | 0.9(0.7-1.0) |
| **Treatments** |  |  |  |  |  |  |  |  |  |
| WD treatment (Reference level: No treatment) |  |  |  |  |  |  |  |  |  |
| D-penicillamine | 10(3.2%) | 32(1.2%) | 1.9(1.0-3.7) | 5(2.0%) | 48(1.6%) | 1.0(0.4-2.4) | 3(0.5%) | 58(2.0%) | 0.2(0.1-0.7) |
| Trientine | 15(4.8%) | 55(2.1%) | 2.0(1.2-3.3) | 8(3.1%) | 81(2.7%) | 1.0(0.5-2.1) | 4(0.7%) | 96(3.2%) | 0.2(0.1-0.5) |
| Zinc | 14(4.5%) | 220(8.5%) | 0.5(0.3-0.9) | 16(6.3%) | 248(8.3%) | 0.7(0.4-1.2) | 80(13.2%) | 245(8.3%) | 1.4(1.1-1.8) |
| Loop diuretics | 34(11.0%) | 230(8.9%) | 1.5(1.0-2.1) | 65(25.5%) | 289(9.6%) | 3.5(2.7-4.7) | 175(29.0%) | 303(10.3%) | 3.8(3.2-4.6) |
| **Health Care Utilization** |  |  |  |  |  |  |  |  |  |
| Number of outpatient visits | 1.8 ± 1.2 | 1.9 ± 1.1 | 1.0(0.9-1.1) | 2.0 ± 1.1 | 1.9 ± 1.1 | 1.1(1.0-1.3) | 2.1 ± 1.1 | 1.8 ± 1.1 | 1.3(1.2-1.4) |
| Number of inpatient visits | 0.4 ± 0.6 | 0.4 ± 0.6 | 1.0(0.8-1.2) | 0.7 ± 0.7 | 0.4 ± 0.6 | 1.9(1.6-2.2) | 0.8 ± 0.7 | 0.4 ± 0.6 | 2.2(2.0-2.4) |
| Length of hospital stay | 0.6 ± 1.0 | 0.7 ± 1.1 | 1.0(0.9-1.1) | 1.2 ± 1.2 | 0.7 ± 1.1 | 1.4(1.3-1.5) | 1.4 ± 1.3 | 0.7 ± 1.1 | 1.6(1.5-1.7) |
| Number of prescriptions | 1.9 ± 1.1 | 1.9 ± 1.1 | 1.1(1.0-1.2) | 2.2 ± 1.0 | 1.9 ± 1.1 | 1.3(1.2-1.5) | 2.1 ± 1.1 | 2.0 ± 1.1 | 1.3(1.2-1.4) |

In the cohort for liver failure, 3,251 patients had WD diagnosis, among whom 255 developed liver failure. Compared with patients without liver failure, liver failure patients had more males (46.7% vs. 43.7%); were more likely to have a history of cirrhosis (44.3% vs. 7.2%), alcoholism (27.8% vs. 6.1%), ascites (31.8% vs. 5.5%), liver fibrosis (32.2% vs. 5.8%), chronic hepatitis (12.5% vs. 4.4%), diabetes (31.0% vs. 20.1%), hepatic steatosis (19.2% vs. 10.5%), and renal dysfunction (25.5% vs. 17.6%); and had a higher proportion of having elevated bilirubin levels (27.1% vs. 5.4%), elevated AST levels (22.0% vs. 6.2%), and low albumin levels (38.4% vs. 15.4%).

In the cohort for death, 3,559 patients had WD diagnosis, among whom 604 patients died. Compared with patients who survived, patients who died were older (62.1 vs 46.7 years); had more males (50.3% vs 42.8%); had a higher proportion of having cancer (23.0% vs 8.9%), renal/kidney failure (33.1% vs. 12.9%), cirrhosis (21.5% vs 12.9%), alcoholism (16.9% vs. 9.6%), hypertension (63.7% vs 37.3%), anemia (57.3% vs. 35.0%), presence of both neurologic and hepatic conditions (21.4% vs. 12.7%), coronary artery disease (27.6% vs. 10.2%), gait disturbance (17.4% vs. 10.2%), dysphagia (10.1% vs. 6.1%), cardiovascular disease (38.1% vs. 17.3%), and chronic kidney disease (38.1% vs. 18.2%). Additionally, they were more likely to use loop diuretics (29.0% vs. 10.3%); have elevated bilirubin levels (16.4% vs. 8.8%), elevated creatinine levels (16.9% vs. 4.7%), low albumin levels (37.9% vs. 16.3%), elevated WBC levels (16.6% vs. 5.5%), elevated AST levels (12.1% vs. 8.6%); and have more inpatient visits and stay longer in the hospital.

### Ensemble Features

The top 20 most important features determined by each model for each study outcome is illustrated in Supplemental Figure 1. The weighted average ranking was calculated for each feature retained by each model. The resulting top 50 ensemble features incorporating the importance ranking across three models for each outcome were reviewed by the clinical specialists and the final ensemble features for each outcome are presented in Table 2. Overall, the increased age at WD diagnosis, history of alcoholism, elevated AST levels, and bilirubin levels within 3 months of WD diagnosis, and presence of neurologic and hepatic conditions were the most common predictive features for progression to cirrhosis, liver failure, and death. Chronic hepatitis, hepatic steatosis, and diabetes were predictive for cirrhosis and liver failure while ascites, albumin levels, anemia, and history of cirrhosis were associated with liver failure and death. The crude hazard ratios of the final ensemble features are reported in Table 1.

**Table 2.** Final Ensemble Features for Individual Risk Prediction.

| **Features for Cirrhosis** | **Features for Liver Failure** | **Features for Death** |
| --- | --- | --- |
| Chronic hepatitis | Cirrhosis | Loop diuretics |
| Alcoholism | Alcoholism | Ethnicity |
| Bilirubin level* | Ascites | Disorders of fluid, electrolyte, and acid-base balance |
| AST level* | Bilirubin level* | Cancer (except non melanoma skin cancer) |
| Acute hepatitis | AST level* | Bilirubin level |
| Neurologic or hepatic conditions | Albumin level* | Ascites |
| Age at WD diagnosis | Anemia | Creatinine level |
| Elevation of lactic acid dehydrogenase level | Parkinsons disease | Age at WD diagnosis |
| Hepatic steatosis | Liver fibrosis | Albumin level |
| Diabetes | Psychosis | Renal/kidney failure |
| Obesity | Chronic hepatitis | Cirrhosis |
| Number of prescriptions | Neurologic or hepatic conditions | Number of inpatient visits |
| WBC level* | Personality disorders | Hepatic encephalopathy |
| Kayser-Fleischer rings | Diabetes | Alcoholism |
| Choreiform movements | Hepatic steatosis | WBC level* |
| WD treatment | Renal dysfunction | Hypertension |
| Coronary artery disease |  | AST level* |
|  |  | Anemia |
|  |  | Neurologic or hepatic conditions |
|  |  | Length of hospital stay |
|  |  | Coronary artery disease |
|  |  | Gait disturbance |
|  |  | Male |
|  |  | Dysphagia |
|  |  | Cardiovascular disease |
|  |  | Chronic kidney disease |

\*Within 3 months of WD diagnosis

### Model performance and Patient-level Prediction

Table 3 reveals that XGB model improved the discrimination ability with a favorable C-statistics value of 0.73 (vs 0.71 for RSF) for cirrhosis; 0.78 (vs 0.77for RSF) for liver failure; and 0.80 for both XGB and RSF for death. Individual patient risks were then assessed for progression to cirrhosis, liver failure, and death using the XGB model with the ensemble features from Table 2. The dynamic AUC was 0.78 at Year 1, 0.74 at Year 2, 0.72 at Year 3 and 0.72 at Year 5 for cirrhosis; 0.82 at Year 1, 0.78 at Year 2, and 0.77 at both Year 3 and Year 5 for liver failure; 0.81 at Year 1, 0.83 at Year 2, and 0.82 at both Year 3 and Year 5 for death (Figure 1). Figure 2 shows the individual survival probability of each outcome at 1, 2, 3, 5 years in the validation set.

**Figure 1.**
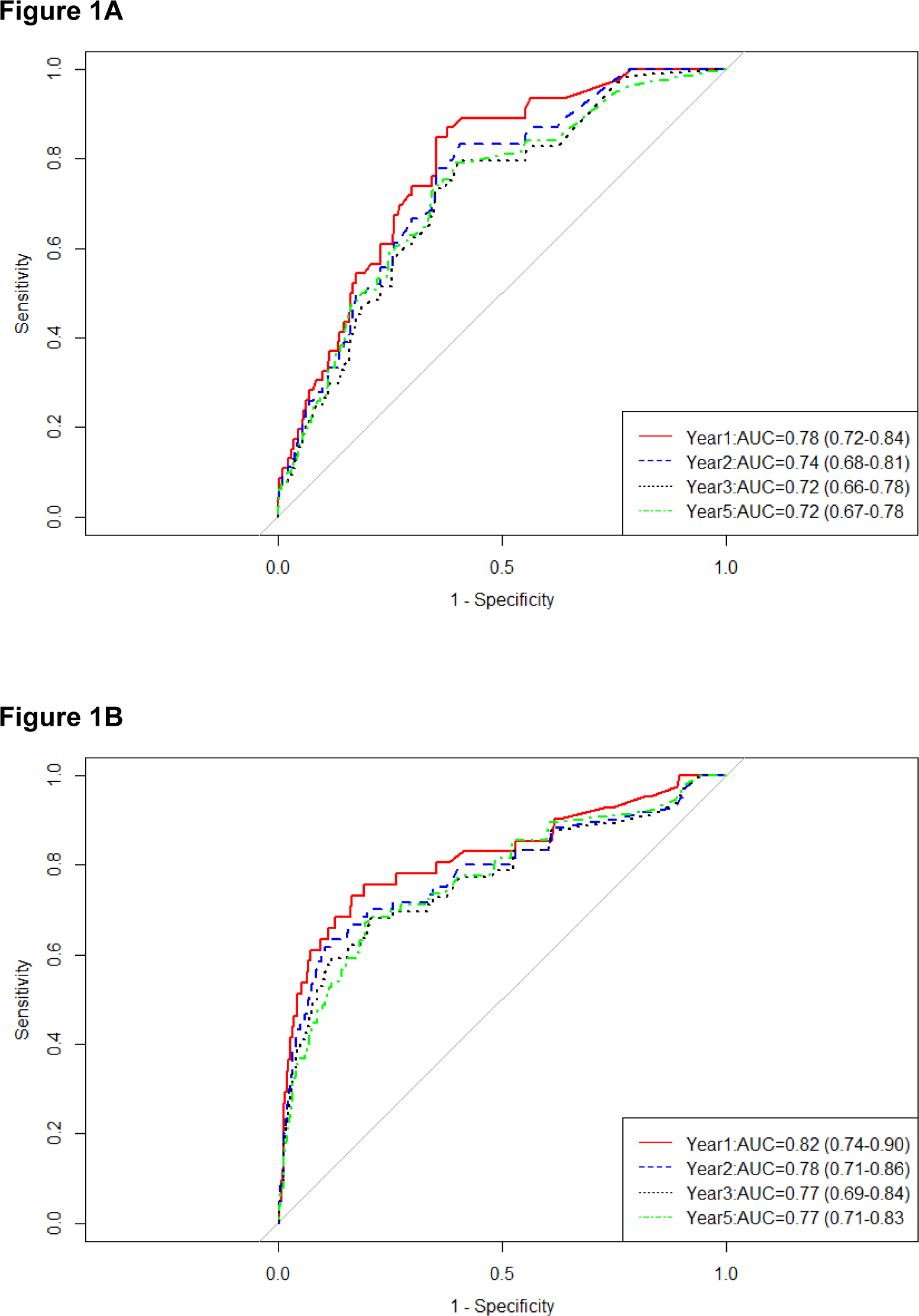
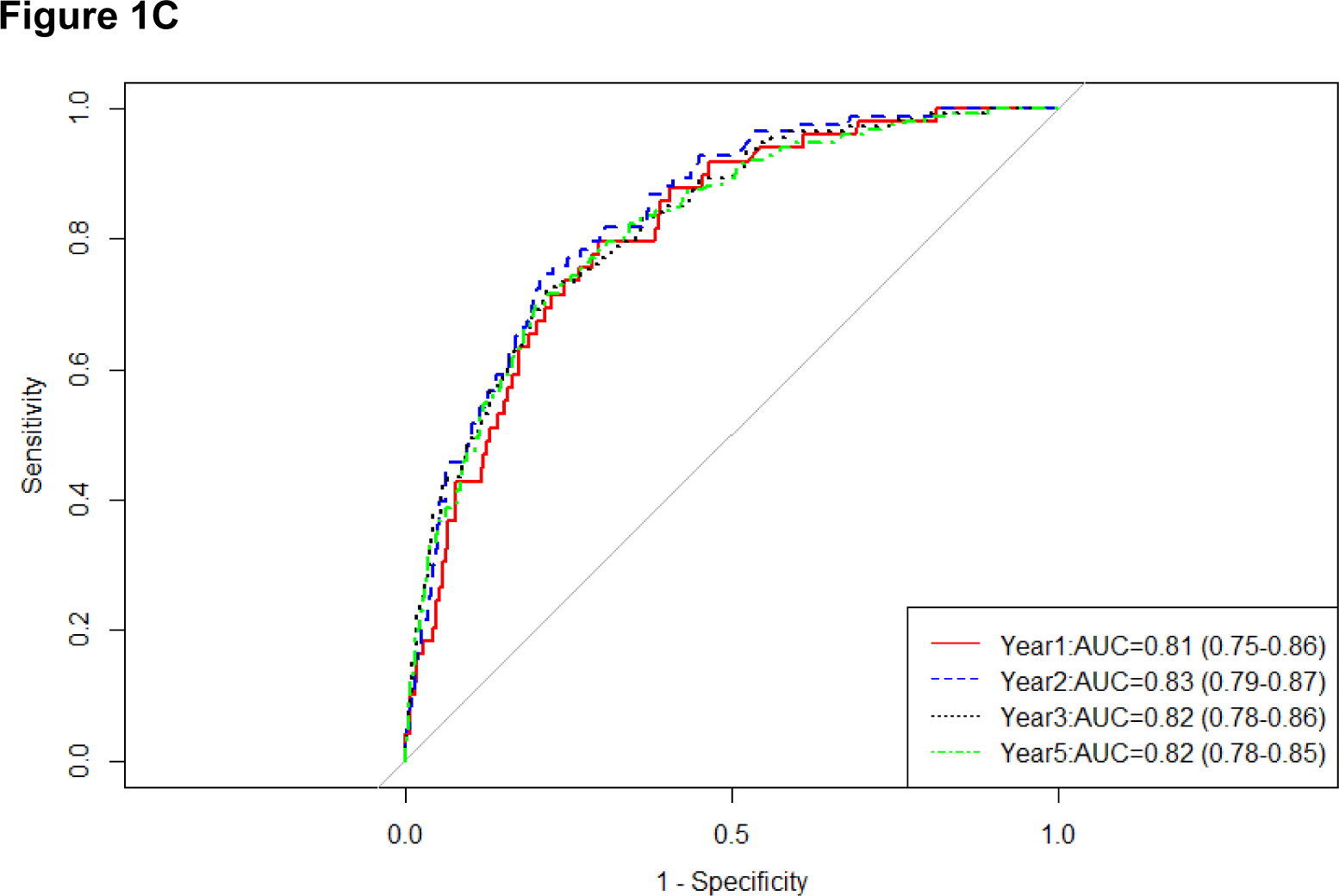
Dynamic AUCs using XGB model for Individual-level Prediction (A. Cirrhosis; B. Liver Failure; C. Death)

**Figure 2.**
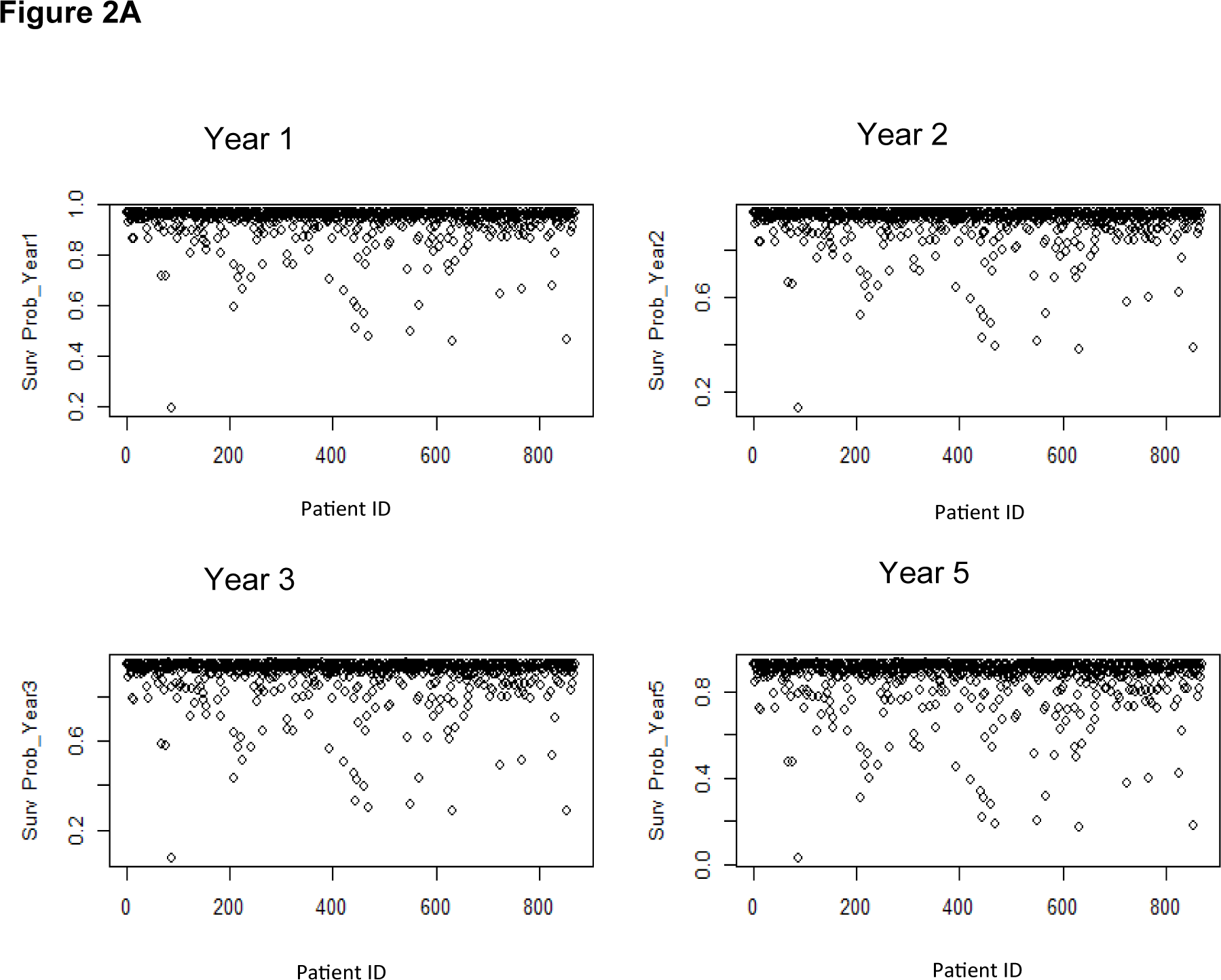
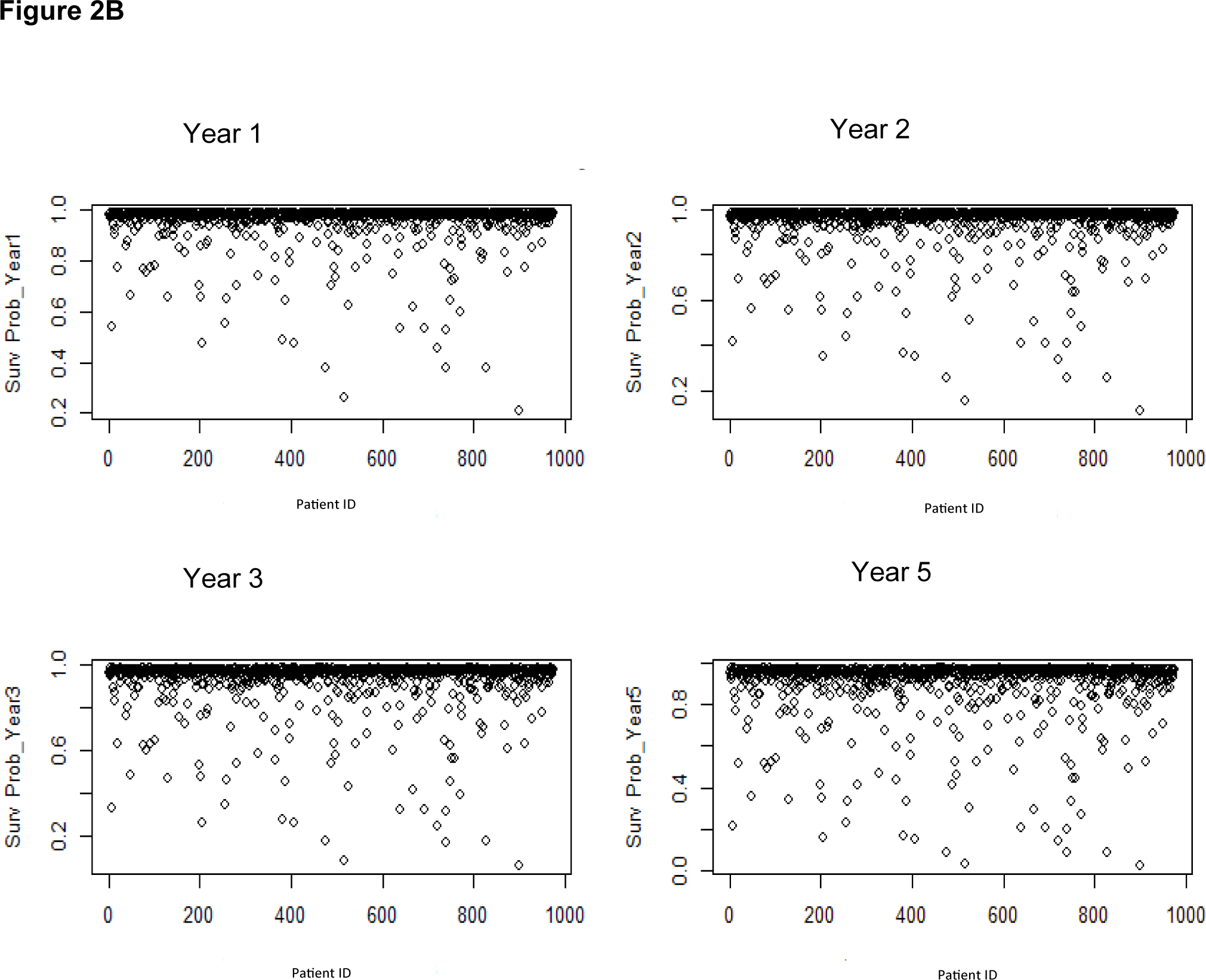
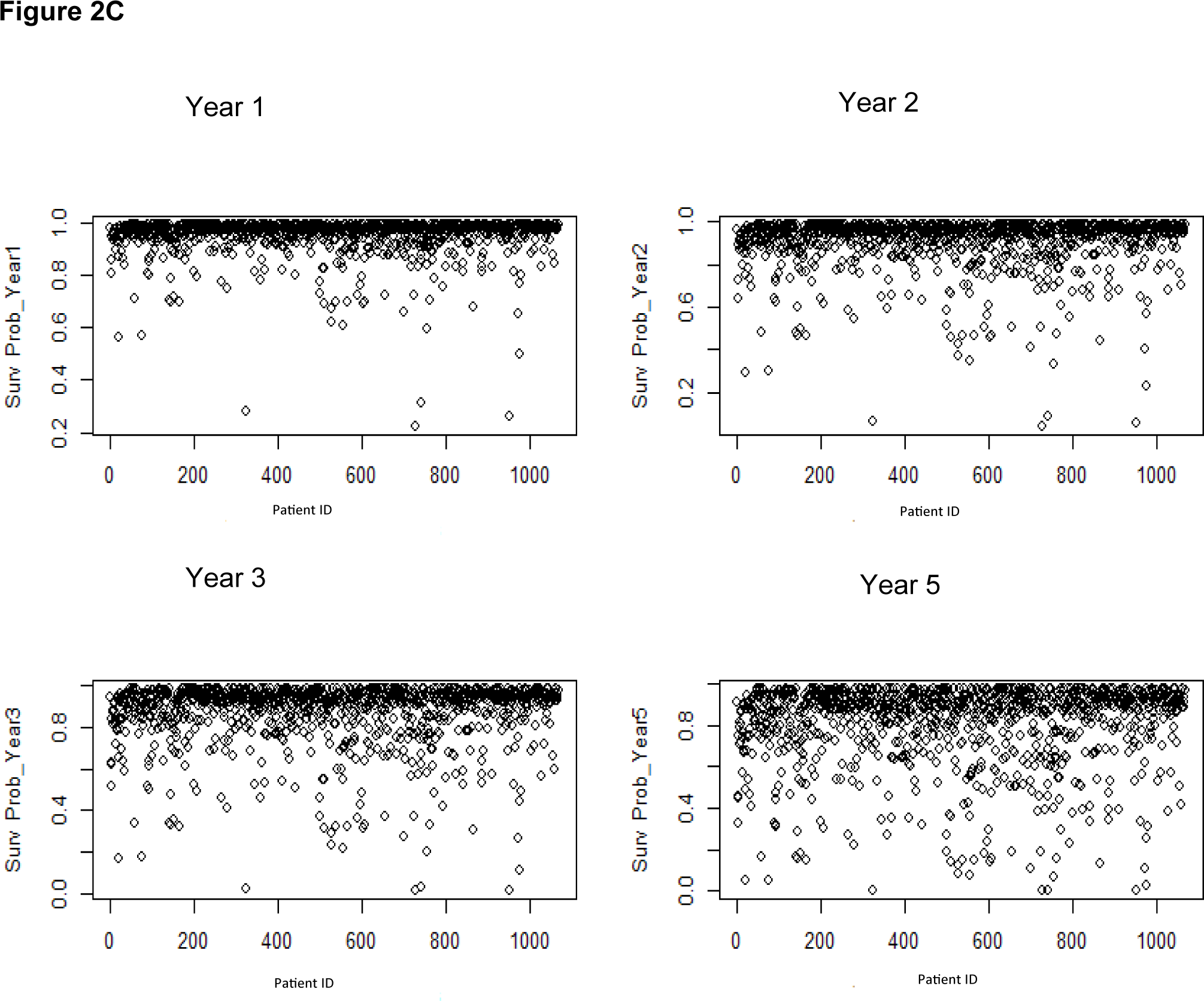
Prediction of Individual Survival Probability (A. Cirrhosis; B. Liver Failure; C. Death)

**Table 3.** C-index for XGB and RSF.

| **Model** | **Cirrhosis** | **Liver Failure** | **Death** |
| --- | --- | --- | --- |
| **RSF** | 0.71 | 0.77 | 0.8 |
| **XGB** | 0.73 | 0.78 | 0.8 |

## Discussion

To our best knowledge, this is the first study to predict WD disease progression using a large US EHR database. In this study, we identified 17 independent predictors for cirrhosis, 16 for liver failure and 26 for all-cause of death among WD patients. This study indicates that checking patient’s medical history of alcoholism, neurologic and hepatic conditions, diabetes and monitoring the laboratory indicators (e.g. AST, bilirubin, creatinine, WBC, albumin) can predict patients at high risk of progression to cirrhosis and liver failure. In addition, age at diagnosis, history of cancer, kidney dysfunction, cirrhosis, hepatic encephalopathy, and cardiovascular disease and use of loop diuretics can predict patient’s all-cause mortality after WD diagnosis. This study also predicts individual survival probability of cirrhosis, liver failure and death at 1, 2, 3, and 5 years since WD diagnosis, which identifies patients with specific profiles who might be at high risk of progression to these outcomes of interest.

There are few studies predicting WD progression. Chen et al ^16^ using XGB to predict liver cirrhosis in WD found that platelet large cell count (P-LCC), red cell distribution width CV (RDW-CV), serum ceruloplasmin, age at diagnosis, and mean corpuscular volume (MCV) were the top five important predictors of liver cirrhosis; in contrast, the top five important features for liver cirrhosis in our study were chronic hepatitis, alcoholism, bilirubin level, AST level and acute hepatitis. Also, Chen et al found D-penicillamine treatment was inversely associated with cirrhosis while we found zinc treatment is more relevant. Consistently, age at diagnosis and WBC level were important features in both studies. The discrepancies between both studies could be explained by the following reasons. First, our study was a retrospective cohort study with a larger sample size (>3,000 WD patients) using machine learning with time-to-event analysis, while Chen et al conducted a case-control study of 346 WD patients based on the analysis without considering the length of time until the occurrence of cirrhosis, which is critical for predicting disease progression. Second, we leveraged a variety of features (about 180 features) including demographics, comorbidities, medical history, lab results, treatment, and health care utilization that are potentially associated with WD progression, while Chen et al mainly focused on the blood-based lab results at the time of WD diagnosis. Although lab results are important predictors, they are considered short-term indicators that could be affected by treatments or other supportive care. Third, feature selection in our study was not driven by a single model, but ensembled from three different models (LASSO, RSF, and XGB), incorporating the review feedbacks from clinical specialists. In Chen et al’s study, the important predictors were identified by XGB only and the AUC of the XGB was 0.79 given a number of 37 features identified. Instead, the dynamic AUC for cirrhosis in our study was 0.78 at Year 1, 0.74 at Year 3, and 0.72 at Year 5 with only 17 independent features. Fourth, overfitting seems to be an issue in Chen et al’s study (an AUC of 0.9998 in the training set and 0.7873 in the testing set), either because of the small sample, narrow variety of features, or lack of bootstrap sampling or cross-validation. In a study of Chaudhuri et al^17^ with a sample of 52 WD patients, gait and age at diagnosis identified by random forest (accuracy was 0.58) were important features for ATP7B gene mutations. In presenting study, these two features were identified to be strong predictors of all-cause of death in WD patients. Zinc therapy has been reported to have similar effects to D-penicillamine on preventing or reducing hepatic or neurological WD symptoms. Zinc therapy has also been shown to be safer and inversely impact mortality.^18^ In our study, Zinc therapy was negatively associated with cirrhosis (crude hazard ratio: 0.51 [0.3-0.88]). Devarbhavi et al^19^ found hepatic encephalopathy and total bilirubin were significantly associated with mortality among WD patients, consistent with our results.

Prediction of disease progression is a critical challenge. Meeting this challenge would allow appropriate planning for individualized care (e.g. identifying patients at high risk for rapid progression). More importantly, individual-level prediction would facilitate the efficient execution of clinical trials by properly deciding the inclusion and exclusion criteria, recruiting patients that are at high risk and who are most beneficial for the potential treatment. Kanwal et al developed and compared 3 machine learning algorithms to identify predictors and predict individuals with cirrhosis at high risk of mortality at 1, 2, and 3 years (AUCs, 0.78, 0.76, and 0.71, respectively).^20^ In this study, we developed machine learning models to predict individual risk of progression to cirrhosis, liver failure and death. The dynamic AUCs (>0.7) at 1, 2, 3 and 5 years provide insight into how performance can change over time. For cirrhosis and liver failure, the performances at Year 3 and Year 5 slightly decrease compared with Year 1 and Year 2, however for death, the performances are consistent over time. This can be considered natural as in the early years, it may be more powerful to predict an earlier future, which cirrhosis and liver failure tended to develop earlier, while all-cause of death may happen in a later year. In this study, our results suggested that XGB had a slight superior performance compared with RSF, consistent with Moncada-Torres et al’s study.^21^ These two models are both ensemble methods but handling the predictions differently. RSF builds classification trees based on out-of-bag data and aggregates the results from all the trees.^7, 22^ XBG iteratively train an ensemble of decision trees, with each iteration using the error residuals of the previous model to fit the next model.^15^ Random forest utilizes bagging to minimize the variance and overfitting, while XGB minimizes the bias and underfitting via bagging and boosting.^8, 10^

The study results should be interpreted with these limitations. First, our study was a retrospective cohort study that identified the first-recorded WD diagnosis during the study period using EHR data. Although we removed patients with prior history of WD or outcomes of interest, it is possible that WD diagnosis or outcomes of interests might have occurred at an earlier time than captured in this study, either because took place outside of the study period or outside of the provider network. Additionally, a diagnosis code may not actually indicate the presence of a disease in EHR data, as the diagnosis codes could be incorrectly coded or used as a rule-out criterion. Uncertainty of the first diagnosis date of WD and its first occurrence of cirrhosis and liver failure could lead to the deviation of the temporality. Second, the missing data in this study were classified in a separate category, especially in the lab results. Multiple imputation or imputation using machine learning could provide a better estimation. Also, a significant amount of missing data in the lab results limits the power of prediction. The copper level and INR were missing about 70-90%. These two indicators are strongly associated with WD disease progression.^24^ A well-capture of these two indicators would improve the model prediction.

In conclusion, this study identified the most influential clinical predictors and assessed individual risk of WD progression using machine learning. As clinical evidence evolves to include more real-world data, results from machine learning prognostic models will increase understanding of disease natural history and etiology and may help improve clinical trial design and guide individualized clinical care. Future studies with large sample are encouraged to validate the results presented in this study.

## Supplemental Materials

### Data Preprocessing

There were a wide range of potential predictors (∼180) for selection on a priori hypothesis based on the literature and clinical knowledge, including demographics, comorbidities, lab results and health care utilization as well as the 30 most frequent diagnoses, procedures, and medications. Demographics included age, gender, race, ethnicity, and region. Comorbidities identified from the 12 months baseline period prior to the WD diagnosis included neurologic or hepatic conditions as well as other ∼60 diagnoses related to alcoholism, chronic or acute hepatitis, diabetes, obesity, cardiovascular disease, liver disease, renal disease, hypertension, depression and so on. Lab results were also extracted in terms of serum levels of bilirubin, albumin, creatinine, alanine aminotransferase (ALT), and aspartate aminotransferase (AST), and white cell counts within 3 months before and after the WD diagnosis. All laboratory results were categorized as lower than normal, normal, and above normal. Health care utilization was defined by the number of inpatient visits, outpatient visits, length of hospital stay and number of prescriptions during 12 months prior to the WD diagnosis. Dummy variables were used for categorical variables. Missing data (if applicable) were coded as a separate category for the particular variable.

### Ensemble Selection

LASSO is powerful in feature selection because it automatically selects crucial features by penalizing the magnitude of features coefficients while minimizing the error between predictions and actual observations.^11^ A 20-fold cross validation method was applied to find the regularization parameter lambda which gave the minimum mean cross-validated errors. Predictors with nonzero coefficients in the LASSO regression model were chosen. RSF and XGB had been widely applied in identifying critical features .^6, 12^ A combination of hyperparameters with 500 trees, 10 splits and a node size of 10 were used to build RSF model and identify the important features. The hyperparameters of XGB included 1000 rounds of iterations, 5-fold cross validation, and a learning rate of 0.06.

### Modeling

The R package of randomForestSRC was used to construct the RSF model using the final ensemble features for each outcome. Classification trees were built on bootstrap samples from the training set. Each bootstrap sample excludes on average 37% of the training data, called out-of-bag data (OOB data). For each bootstrap sample, a survival tree is grown based on a splitting criterion. For each tree, a cumulative hazard function (CHF) is calculated. The CHF from all trees were averaged to obtain the ensemble CHF. The prediction performance is then calculated using OOB data in the training set and later in the validation set. The number of trees grown, optimal node size and number of candidate features selected at each split were tuned to find the best model performance.

The R package of xgboost was used to build the XGB model using the final ensemble features for each outcome. In the XGB prediction model, the hyperparameters were tuned via 1000 iterations and 5 cross-validation to get the best accuracy and avoid overfitting. Different from the RSF that aggregates the results from all the trees, XGB estimated the target function by adapting the gradient boosting with maximum partial likelihood in the function space. At each iteration, the target function is updated on the direction of its negative gradient fitted through a regression-tree based on a random subsample of the training set. The final estimated function is chosen at the iteration when optimal out-of-bag prediction performance is achieved.^15^

**Figure 1A.**
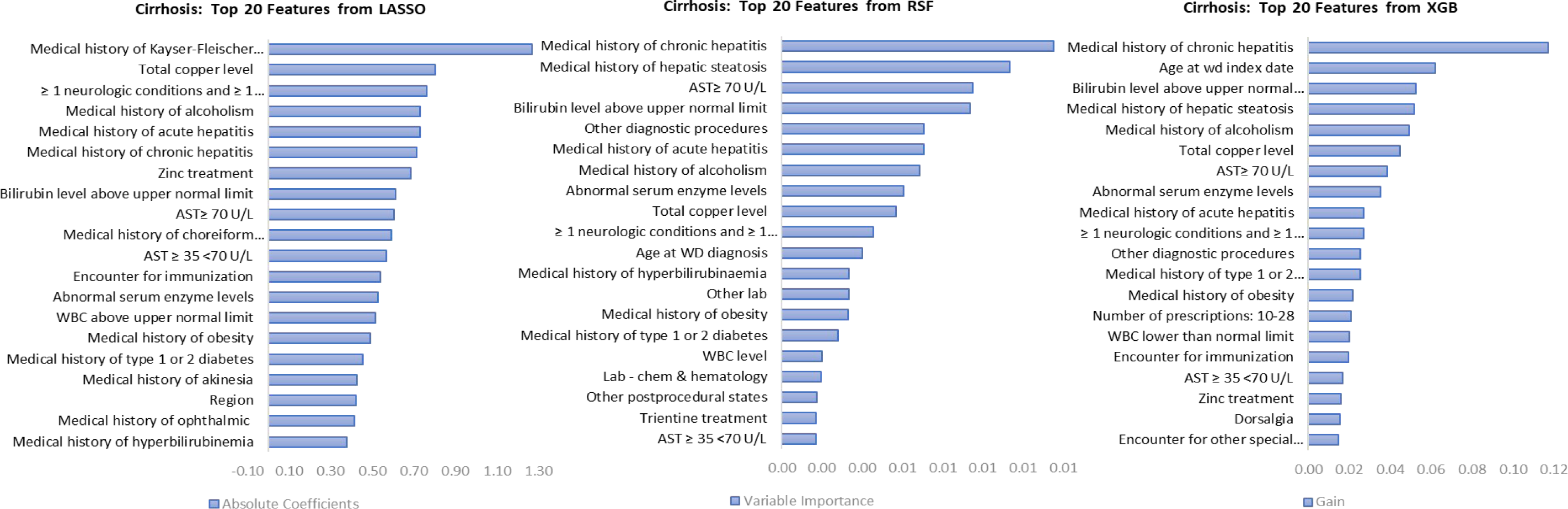
Top 20 Most Important Features for Cirrhosis.

**Figure 1B.**
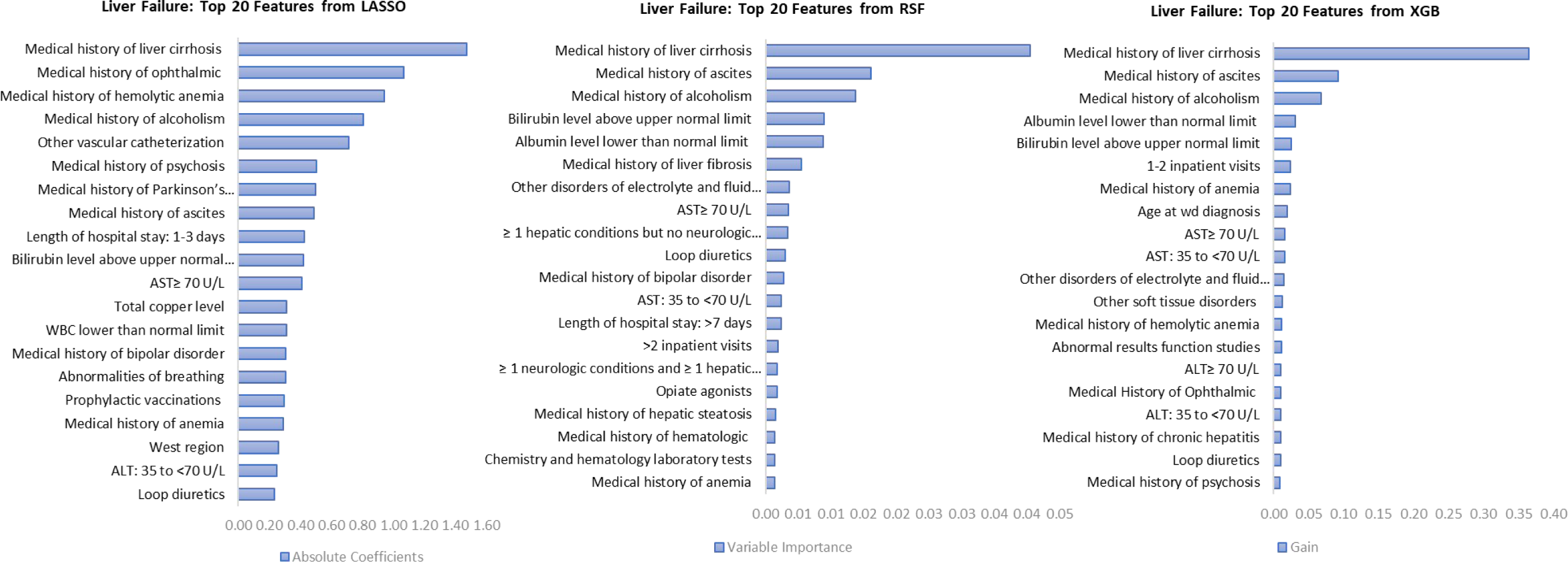
Top 20 Most Important Features for Liver Failure.

**Figure 1C.**
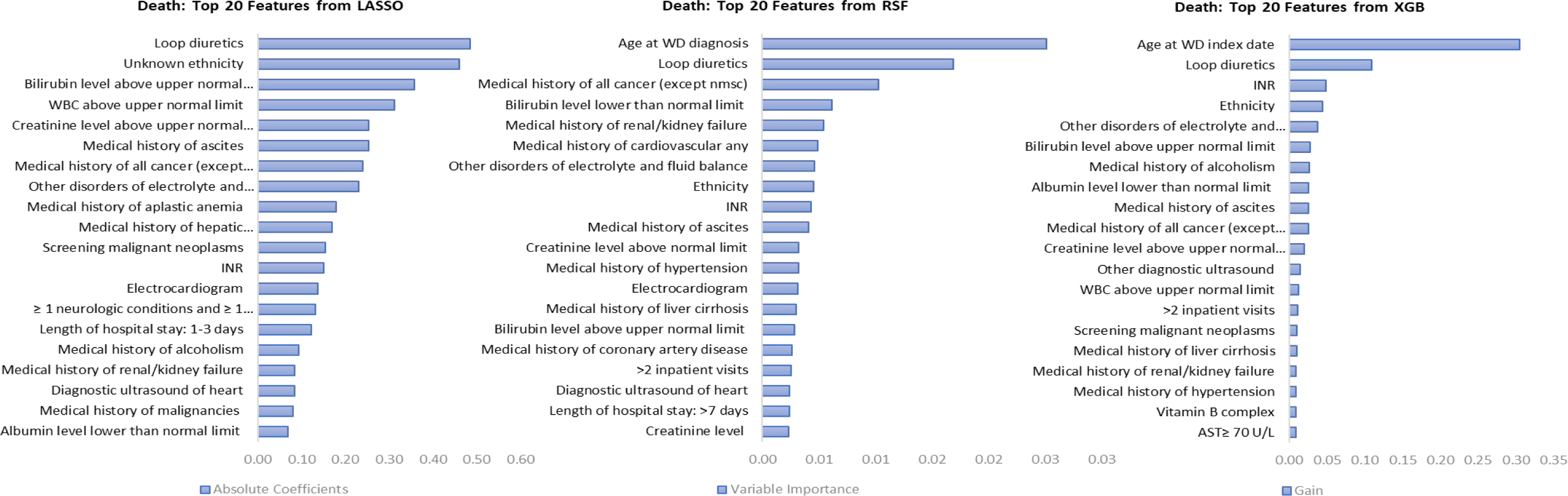
Top 20 Most Important Features for death.

## References

1. Chang IJ, Hahn SH. The genetics of Wilson disease. Handb Clin Neurol 2017;142:19–34.

2. Pfeiffenberger J, Mogler C, Gotthardt DN, et al. Hepatobiliary malignancies in Wilson disease. Liver Int 2015;35(5):1615–22.

3. Boga S, Ala A, Schilsky ML. Hepatic features of Wilson disease. Handb Clin Neurol 2017;142:91–99.

4. Omurlua IK TM, Tokatlib F. The comparisons of random survival forests and Cox regression analysis with simulation and an application related to breast cancer. Expert Syst Appl, 36, 8582–8 2009.

5. Miao F, Cai YP, Zhang YX, Li Y, Zhang YT. Risk Prediction of One-Year Mortality in Patients with Cardiac Arrhythmias Using Random Survival Forest. Comput Math Methods Med 2015;2015:303250.

6. Spooner A, Chen E, Sowmya A, et al. A comparison of machine learning methods for survival analysis of high-dimensional clinical data for dementia prediction. Sci Rep 2020;10(1):20410.

7. Ishwaran HK, UB; Blackstone, EH; Lauer, MS. Random survival forests. . Ann Appl Stat 2 (3) 841–860 2008.

8. Wang H, Zhou L. Random survival forest with space extensions for censored data. Artif Intell Med 2017;79:52–61.

9. C. CTG. XGBoost: a scalable tree boosting system. In: The 22nd ACM SIGKDD International Conference on Knowledge Discovery and Data Mining (KDD ’16) Association for Computing Machinery, New York, NY, USA, pp 785–794 2016.

10. Jiang J, Pan H, Li M, Qian B, Lin X, Fan S. Predictive model for the 5-year survival status of osteosarcoma patients based on the SEER database and XGBoost algorithm. Sci Rep 2021;11(1):5542.

11. Muthukrishnan RR, R. LASSO: A feature selection technique in predictive modeling for machine learning. 2016 IEEE International Conference on Advances in Computer Applications (ICACA) 2016.

12. Li MX, Sun XM, Cheng WG, et al. Using a machine learning approach to identify key prognostic molecules for esophageal squamous cell carcinoma. BMC Cancer 2021;21(1):906.

13. Ishwaran H, Gerds TA, Kogalur UB, Moore RD, Gange SJ, Lau BM. Random survival forests for competing risks. Biostatistics 2014;15(4):757–73.

14. H WjXjZCPYW. An ensemble feature selection method for high-dimensional data based on sort aggregation. Systems Science & Control Engineering 2019;7(7):32–39.

15. Friedman JH. Contrast trees and distribution boosting. Proc Natl Acad Sci U S A 2020;117(35):21175–21184.

16. Chen K, Wan Y, Mao J, Lai Y, Zhuo-Ma G, Hong P. Liver cirrhosis prediction for patients with Wilson disease based on machine learning: a case-control study from southwest China. Eur J Gastroenterol Hepatol 2022;34(10):1067–1073.

17. Chaudhuri J, Biswas S, Gangopadhyay G, et al. Correlation of ATP7B gene mutations with clinical phenotype and radiological features in Indian Wilson disease patients. Acta Neurol Belg 2022;122(1):181–190.

18. Appenzeller-Herzog C, Mathes T, Heeres MLS, Weiss KH, Houwen RHJ, Ewald H. Comparative effectiveness of common therapies for Wilson disease: A systematic review and meta-analysis of controlled studies. Liver Int 2019;39(11):2136–2152.

19. Devarbhavi H, Singh R, Adarsh CK, Sheth K, Kiran R, Patil M. Factors that predict mortality in children with Wilson disease associated acute liver failure and comparison of Wilson disease specific prognostic indices. J Gastroenterol Hepatol 2014;29(2):380–6.

20. Kanwal F, Taylor TJ, Kramer JR, et al. Development, Validation, and Evaluation of a Simple Machine Learning Model to Predict Cirrhosis Mortality. JAMA Netw Open 2020;3(11):e2023780.

21. Moncada-Torres A, van Maaren MC, Hendriks MssP, Siesling S, Geleijnse G. Explainable machine learning can outperform Cox regression predictions and provide insights in breast cancer survival. Sci Rep 2021;11(1):6968.

22. Wang H, Li G. A Selective Review on Random Survival Forests for High Dimensional Data. Quant Biosci 2017;36(2):85–96.

23. Ferenci P, Czlonkowska A, Merle U, et al. Late-onset Wilson’s disease. Gastroenterology 2007;132(4):1294–8.

24. Schilsky ML. Wilson disease: Clinical manifestations, diagnosis, and treatment. Clin Liver Dis (Hoboken) 2014;3(5):104–107.

